# Short-term mental health sequelae of bereavement predict long-term physical health decline in older adults: US Health and Retirement Study Analysis

**DOI:** 10.1101/19009878

**Authors:** Benjamin W. Domingue, Laramie Duncan, Amal Harrati, Daniel W. Belsky

**Author notes:** Authors can be contacted at BWD; 520 Galvez Mall Stanford CA 94305, DWB; Columbia Aging Center, 722 W 168th Street, New York NY USA 10032. Disclosures: Authors report no financial relationships with commercial interests.

## Abstract

**Objective:** Death of a spouse is a common late-life event with mental- and physical-health sequelae. Whereas mental-health sequelae of spousal death tend to be transient, physical-health sequelae may persist, leading to disability and mortality. Growing evidence linking poor mental health to aging-related disease suggests the hypothesis that transient poor mental health following death of a spouse could be a harbinger of physical health decline. If so, identification of bereavement-related mental health symptoms could provide an opportunity for prevention.

**Methods:** We analyzed data from N=35,103 individuals followed from 1994-2014 in the US Health and Retirement Study (HRS) and identified N=4,629 who were widowed during follow-up. We tested change in mental and physical health from pre-bereavement through the 5-year period following spousal death.

**Results:** Bereaved spouses experienced an immediate increase in depressive symptoms following their spouses’ deaths but the depressive shock attenuated within one year. Bereaved spouses also experienced increases in disability, chronic-disease morbidity, and hospitalization, which grew in magnitude with time since spousal death, especially among older HRS participants. Bereaved spouses were at increased risk of death compared to HRS participants who were not bereaved. The magnitude of depressive symptoms in the immediate aftermath of spousal death predicted physical-health decline and mortality risk over 5 years of follow-up.

**Conclusions:** Bereavement-related depressive symptoms provide an indicator of risk for physical health decline and death in older adults. Screening for depressive symptoms in bereaved older adults may represent an opportunity for intervention to preserve healthy lifespan.

**Plain Language Summary:** The clinical significance of depressive symptoms during bereavement for long-term health is not well understood. We analyzed longitudinal data tracking mental and physical health of more than 4,000 older adults who were widowed during follow-up. Widows and widowers who experienced more severe depressive symptoms immediately following their spouse’s death were at increased risk for incident disability, chronic disease, hospitalization, and mortality over the five years following their spouse’s death. Transient depressive symptoms related to bereavement may provide a clinical indicator of risk for long-term physical health decline in older adults. Findings motivate increased integration of psychiatric assessment in geriatric care.

## INTRODUCTION

Death of a spouse is a common life event for older adults and has significant health consequences. Bereavement is associated with acute increase in symptoms of depression, which manifest right around the time of death and subside within a year (1–7). In contrast to the transience of depressive symptoms, physical-health sequelae of bereavement may be more persistent, as indicated by increased risk of death among the bereaved (8–14). Despite evidence for a link between the two (1), mental- and physical-health sequelae of bereavement have often been studied separately. New data linking mental health symptoms with acceleration of aging-related physical deterioration (15–17) suggest that mental- and physical-health sequelae of bereavement may be connected. If so, this connection would have implications for screening and management of mental-health sequelae of bereavement and potentially for preventive intervention to preserve health in aging.

To test for connection between mental- and physical-health sequelae of bereavement, data are needed that track spouses’ mental and physical health over time, from before a spouse dies through the period of grief and recovery that follows. Three further challenges must be overcome. First, bereavement-related health decline must be distinguished from the typical pattern of health decline that occurs with aging. Second, mental- and physical-health decline may accelerate during the period leading up to spousal death (18,19). Sequelae of the specific shock of bereavement must be distinguished from the health declines during the pre-bereavement period. Third, mental- and physical-health sequelae of bereavement manifest over different time scales; transient mental-health sequelae arise and resolve over months while persistent physical-health sequelae emerge over years. To study links between mental- and physical-health sequelae of bereavement, observations of mental health symptoms during the months following bereavement must be linked with observations of physical health declines occurring over the following years.

We addressed these challenges in analysis of longitudinal data tracking N=35,103 adults aged 51 and older during the period 1994-2014 in the US Health and Retirement Study, a nationally representative study of US older adults (20). During follow-up, we observed bereavement in 4,629 HRS participants. We analyzed data on bereaved participants’ depressive symptoms, disability, chronic disease morbidity, hospitalizations, and mortality. We used exponential-decay analysis to quantify the magnitude of bereavement-related increases in depressive symptoms during the months immediately following spousal death. We then tested if participants who experienced more severe depression in the months following their spouses’ deaths went on to experience more severe declines in physical health over subsequent years.

## METHODS

### Data

The US-based Health and Retirement Study (HRS) is a nationally representative longitudinal study of health and aging ongoing since 1992 (20). HRS includes a nationally-representative sample of adults aged 51 and older and their spouses. We analyzed RAND Corporation’s 2014 HRS data release (v2); these data include N=35,103 participants meeting inclusion criteria (**Supplemental Information A1**). We analyzed data from 1994 on due to changes made to the survey between 1992 and 1994. Summary statistics for HRS health measures are reported in **Supplemental Table S1**.

### Bereavement

HRS records participants’ death dates from the National Death Index and other sources (21). We analyzed death records for pairs of HRS spouses. We calculated time from spousal death as death-date subtracted from HRS-interview-date (**Supplemental Figure S2**). We analyzed data from n=4,629 participants widowed during follow-up and who were interviewed within 3 years of their spouse’s death. Participants who remarried during HRS follow-up (n=971) were excluded. Bereaved participants were more likely to be female (72% vs. 56% in the HRS overall) and to have been born earlier (mean birth-year for HRS participants=1937 as compared to 1930 for our bereaved sample). **Supplemental Table S1** compares the bereaved subsample to the full HRS.

### Mental Health Sequelae of Bereavement: Depressive Symptoms

We quantified mental health sequelae of bereavement using a modified version of the Center for Epidemiologic Studies Depression (CES-D) scale (22,23). Modified CES-D scales have been shown to retain key measurement properties (24,25). Participants gave yes/no responses to eight prompts (e.g., Is everything an effort? Did the respondent feel lonely?) based on their status the week prior to interview. CES-D items are listed in **Supplemental Information A2**.

### Physical-Health Sequelae of Bereavement

We analyzed four physical-health sequelae of bereavement: chronic disease morbidity, disability, hospitalization, and mortality.

#### Chronic disease morbidity

was measured as the count of chronic health conditions that participants reported in response to prompts beginning “Has a doctor ever told you that you had…”. The chronic conditions were high blood pressure, diabetes, cancer, lung disease, heart disease, stroke, and arthritis.

#### Disability

was measured as counts of impairments to Activities of Daily Living (ADLs) and Instrumental Activities of Daily Living (IADLs). We counted all activities for which the participant reported some or more impairment. ADLs included bathing, dressing, eating, getting out of bed, and walking across a room. IADLs included using the phone, handling money, taking medication, shopping, and preparing meals.

#### Hospitalization

was measured as the reported number of hospital stays during the previous 2 years. We coded all values of 10 or greater as 10.

#### Mortality

Date of death was obtained HRS record linkage with the US National Death Index and other sources (22).

### Analysis

We analyzed bereavement sequelae over the five-year interval beginning with the first HRS interview following spousal death. Our analysis aimed to address the three challenges to documenting connection among mental- and physical-health sequelae of bereavement outlined in the introduction. We describe our approach in detail in **Supplemental Information A3**. We outline this approach below.

To address the challenge of distinguishing sequelae of bereavement from typical aging-related health decline, we age-residualized measures of physical-health sequelae prior to analysis based on a model of normative aging. To develop a model for normative aging, we operationalized chronological age as a 5^th^-degree b-spline. We then conducted 100 bootstrap repetitions of the following analysis: We sampled one observation at random from each participant in the HRS sample and regressed the health outcome on the age splines, sex, age-spline-sex interactions, and a series of dummy variables encoding the wave of the observation. We averaged coefficients across the bootstrap repetitions and use these averages to conduct our residualization. The residual values for the health measures carried forward into analysis thus represent deviations of a person’s health from the expectation given their chronological age. **Supplemental Figure S5** shows that age-residualized health measures have no aging-related trends.

We conducted descriptive analysis comparing participants mental and physical health before and after their spouse’s death. For physical health, which showed strong age-patterning in the HRS data, analysis focused on the age-residualized health values described above. We compared health before and after bereavement using person-level fixed-effects regression of health variables on an indicator of whether the observation was taken before or after spousal death (Eqn 3, **Supplemental Information A3**).

To address the challenge of isolating effects of bereavement from health trends preceding spousal death, we used discontinuity models. We fitted person-level fixed-effects regression to repeated measures health data. The person-level fixed-effects regression focuses analysis on within-person change. This model holds constant all characteristics of the individual that do not change over time. To fit the discontinuity model, we conducted fixed-effects regression of health on a variable encoding time, an indicator variable coding whether the observation was taken before or after spousal death, and the interaction of those two variables (**Supplemental Information A3 Eqn 4**). Time values were centered on date of spousal death. The coefficient for the indicator variable estimates immediate change in health at the time of spousal death. The time coefficient estimates the rate of change in health during the time leading up to spousal death. The interaction coefficient estimates the change in health decline occurring with bereavement.

To address the challenge of analyzing mental- and physical health sequelae of bereavement on different time scales, we modeled mental-health sequelae (CES-D depressive symptoms) using an exponential decay model (26). We applied these models to data from the first observation of an individual after their spouses’ death (**Supplemental Information A3 Eqn 1**). The exponential decay model uses CES-D data collected from participants who were interviewed at varying amounts of time from their spouse’s death to estimate a function that describes how depressive symptoms decline with increasing time since spousal death. We previously used this approach to study mental health sequelae of bereavement in the HRS (27). In the current analysis, we used the exponential decay model to calculate residual values for participants that quantified how much more or less depressed they were than would be expected for the average HRS participant observed at the same amount of time following their spouse’s death. To test connections between mental- and physical-health sequelae of bereavement, we inserted these residual values into the discontinuity model. Specifically, we modeled interactions between discontinuity-model terms and the CES-D residual values. The resulting model tests the hypothesis that short-lived depressive symptoms during bereavement predict health decline over the subsequent five years (**Supplemental Information A3 Eqn 5**).

## RESULTS

### Death of a spouse is associated with a transient increase in symptoms of depression

We visualized changes in depressive symptoms occurring following spousal death by plotting LOESS curves (28) of participants’ CES-D scores in the years surrounding their spouses’ deaths (**Figure 1**). HRS participants who lost their spouses experienced a sharp increase in depressive symptoms at the time of death and recovered toward baseline levels of depression within the following year, consistent with patterns observed in previous research (5). In the first two years following their spouses’ deaths, participants scored an average of 0.51 standard-deviations higher on the CES-D (95% CI 0.45-0.56; **Supplemental Table S3, Column 1**) as compared to their CES-D scores from interviews prior to their spouses’ death. By the second interview following their spouse’s death, CES-D scores had returned toward baseline levels; scores were 0.06 standard-deviations higher on the CES-D (95% CI 0.00-0.12; **Supplemental Table S3, Column 3**).

**Figure 1.**
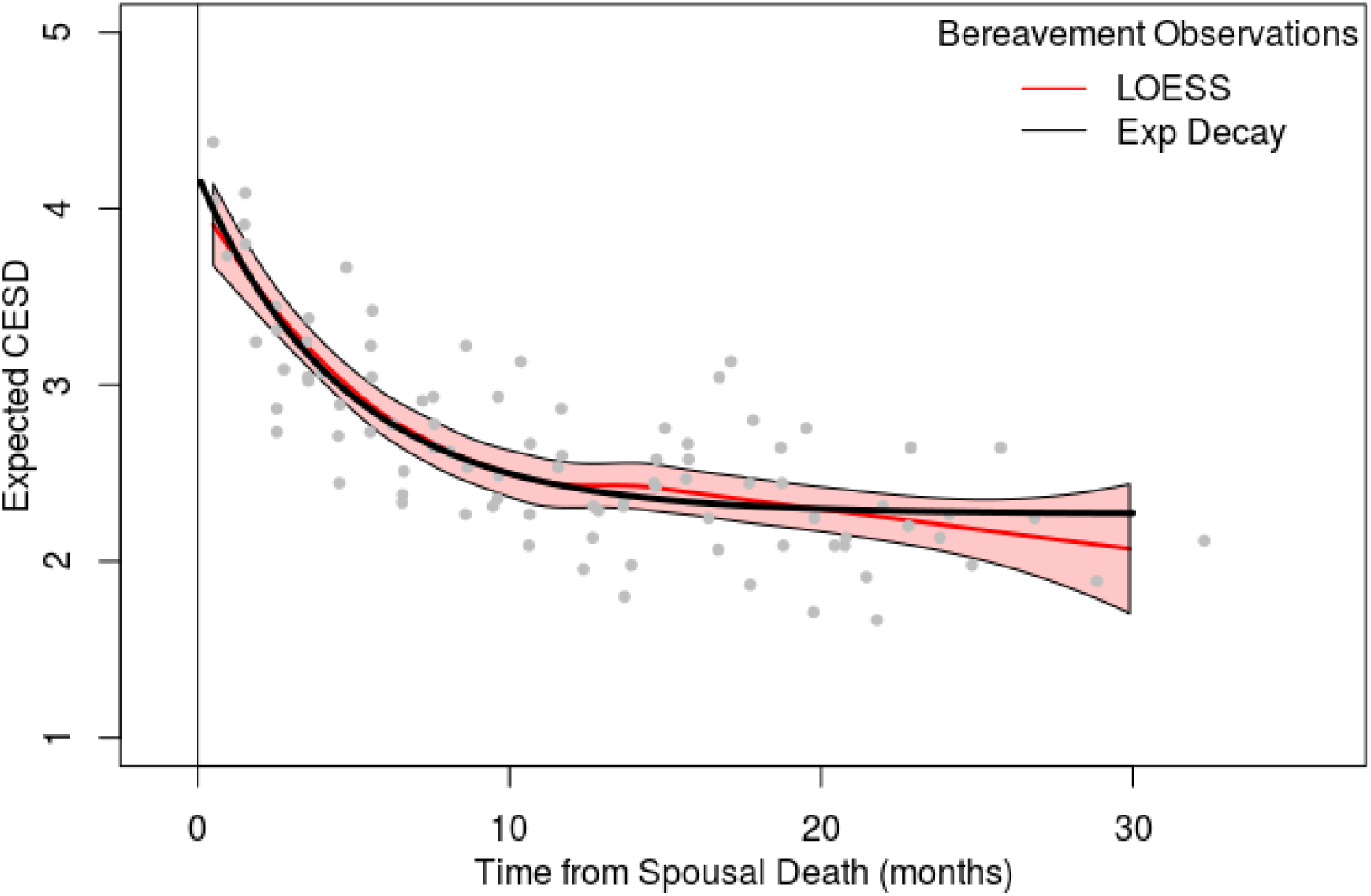
Depressive symptoms after spousal death. The figure shows a binned scatterplot and LOESS-fitted trend-lines illustrating CES-D-measured depressive symptoms during the 3 years before and after spousal death among widowed HRS participants (N=4129). LOESS-based trajectory of depression (in red) for bereavement observations versus fits from exponenential decay models. LOESS and exponential decay estimates are based on original, not binned, data.

To more precisely estimate changes in depressive symptoms following spousal death, we fitted an exponential decay model (26) to CES-D scores from the first interview following spousal death (i.e., Eqn 1, **Supplemental Information A3**). The average participant experienced an increase of 1.92 depressive symptoms immediately following their spouse’s death (95% CI 1.60-2.24) and recovered to near the long-run mean over the next 8 months. Findings were similar for men and women (p-value for test of sex difference = 0.92) and for older and younger participants (p-value for test of age difference = 0.10). Results from all models are reported in **Supplemental Table S2**. Symptoms of loneliness and sadness accounted for most of the bereavement-related increase in CES-D scores (**Supplemental Figure S7 and S8**).

### Death of a spouse is associated with persistent increases in burden of disability, morbidity, and mortality

Parallel to our analysis of depressive symptoms, we found that participants reported increased limitations to instrumental activities of daily living (IADLs), activities of daily living (ADLs), chronic disease morbidity, and hospitalizations within two years of spousal death as compared to levels reported at previous interviews (b=0.02-0.08; **Supplemental Table S3, Column 1**). However, whereas bereaved participants recovered from increased depressive symptoms within a year of their spouses’ deaths, physical health problems worsened over time. Compared with levels measured before their spouses’ deaths, participants reported larger increases in physical health problems at the second interview following their spouse’s death (24-48 months from spousal death) than they had at the previous interview (b=0.05-0.14; **Supplemental Table S3, Column 3**). Effect-sizes of bereavement-related increase in physical health problems at first and second follow-ups are graphed in **Figure 2**. We then tested if increasing time since spousal death was associated with worsening physical health outcomes using the discontinuity model (**Supplemental Information A3, Eq 4**). Increasing time since spousal death was associated with worsening physical health, over and above aging-related and pre-bereavement trends (**Supplemental Figure S10**); for some outcomes, such as ADLs, this was due primarily to changes for older respondents (**Supplemental Table S4**).

**Figure 2.**
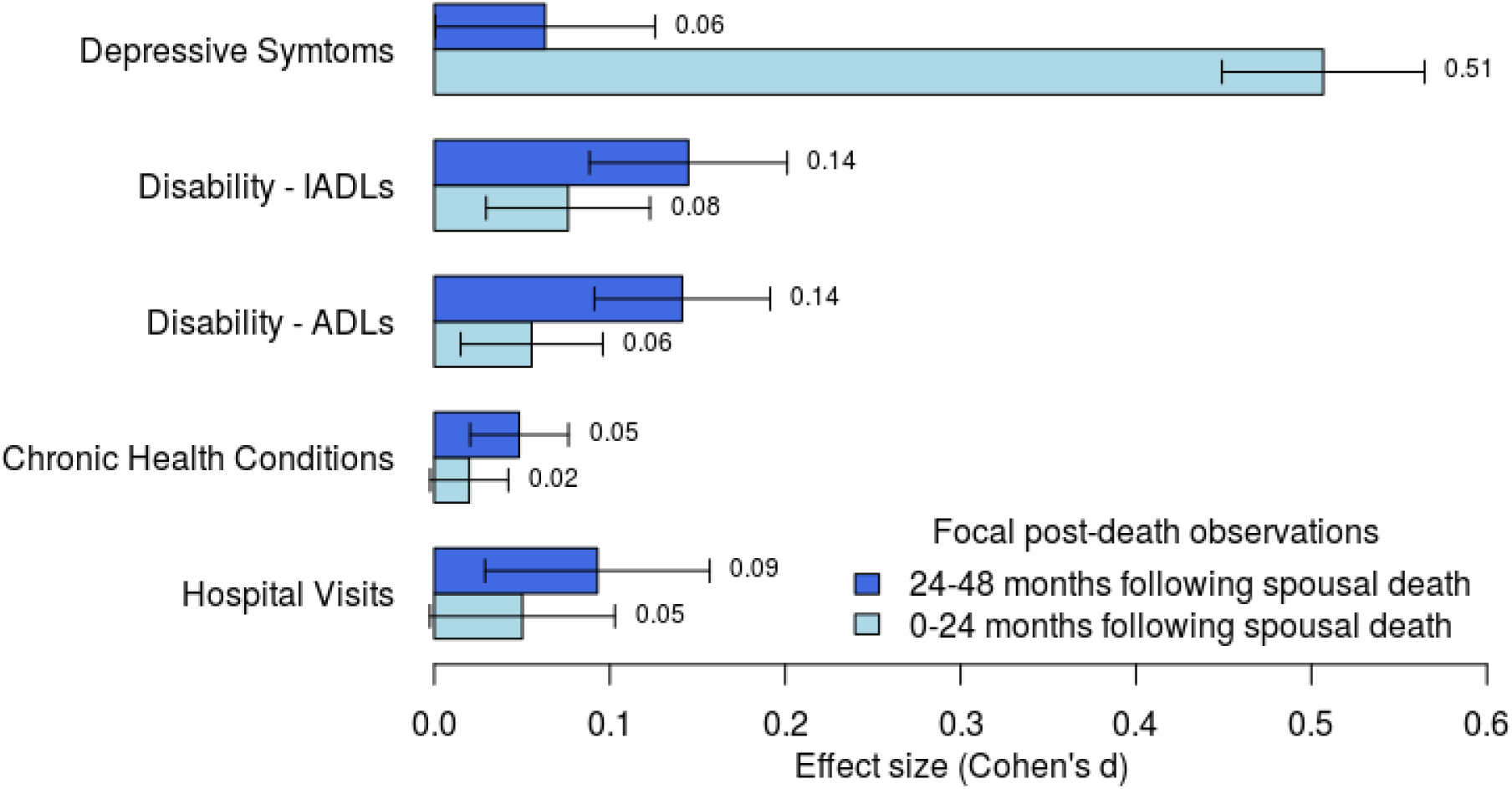
Bereavement-related depressive symptoms decline with time since spousal death, whereas disability and morbidity increase. Figure graphs effect-sizes and 95% confidence intervals for within-person comparisons of CES-D and physical health measures before and after spousal death. Effect-sizes are graphed for comparisons of average levels pre-death to the level at the first observation following spousal death (0-24 months after the death, light blue bars) and at the second observation following spousal death (24-48 months after the death, dark blue bars). Health outcomes were adjusted for aging-related decline and period effects (outcome measures were residualized for age (five splines), sex, interactions between the age-based splines and sex, and HRS observation wave). Effect-sizes are reported in HRS standard deviation units. Results are based on Eqn 3, Supplemental Information A3; full model results are shown in Table S3 (minimum N=11109 observations of N=2865 individuals observed at two HRS observations following spousal death). The figure shows that the increase in depressive symptoms relative to pre-bereavement levels is largest in the first 2 years following spousal death and then declines. In contrast, increases in disability and morbidity grow larger with increasing follow-up time.

Consistent with increase in health problems, bereaved participants were also at increased risk of mortality over the five years of follow-up. Of the bereaved respondents, 21% died during this follow-up interval (**Supplemental Figure S3**). We compared 5y mortality risk for bereaved participants to 5y mortality across all HRS observations. At age 74 years, mortality risk was 2.2 percentage points higher for bereaved participants as compared to others (p<0.001, **Supplemental Table S5**) as compared to participants of the same age and sex who had not experienced bereavement.

### Bereavement-related physical health declines were most pronounced in older HRS participants

Because disability and morbidity are concentrated in older HRS participants (**Supplemental Figure S4B)**, we investigated whether mental- and physical-health sequelae of bereavement differed for older as compared to younger HRS participants. For mental-health sequelae, we found no difference; increases in depressive symptoms following spousal death were similar for younger HRS participants as compared to older HRS participants (**Supplemental Table S2**). In contrast, physical-health sequelae of bereavement were more severe for older HRS participants and less severe for younger ones (see **Supplemental Information C**; results for disability, chronic disease, and hospitalization in **Supplemental Table S3, columns 2 and 4**; results for mortality in **Supplemental Table S5, column 2**).

### HRS participants who experienced more-severe mental-health sequelae immediately following their spouse’s death suffered worse health decline over the following years as compared to participants who experienced less-severe mental-health sequelae

To test connections between transient mental-health sequelae of bereavement and more persistent physical health sequelae, we incorporated results from our exponential decay analysis of depressive symptoms into our discontinuity models testing bereavement effects on physical health. Because physical health sequelae of bereavement were concentrated in older HRS participants, we stratified analysis by age at which the respondent experiences spousal death. We analyzed the role of bereavement-related depression by predicting (based on model results reported in **Supplemental Table S6**) levels of health following spousal death for a hypothetical participant with low levels of bereavement-related depressive symptoms (25^th^ percentile, about 2 fewer symptoms than the norm) versus high levels of depressive symptoms (75^th^ percentile, about 2 more symptoms than the norm). Given that Figure 2 suggests health-related consequences of spousal death accumulate over time, we focused prediction on the moment five years following spousal death. Among older HRS participants (aged 74 and older at time of spousal death, n=2,268), those with more severe depression following their spouse’s death experienced steeper increases in health problems in the subsequent years, primarily in counts of IADLs and ADLs. Differences are illustrated in **Figure 3**. At five years after spousal death, differences in health decline between these groups ranged from b=0.31 for IADLs to b=0.06 for chronic health conditions. Among younger HRS participants (aged 55-74, n=1,861), there were small associations between depressive symptoms and disabilities following bereavement (**Supplemental Figure S11**).

**Figure 3.**
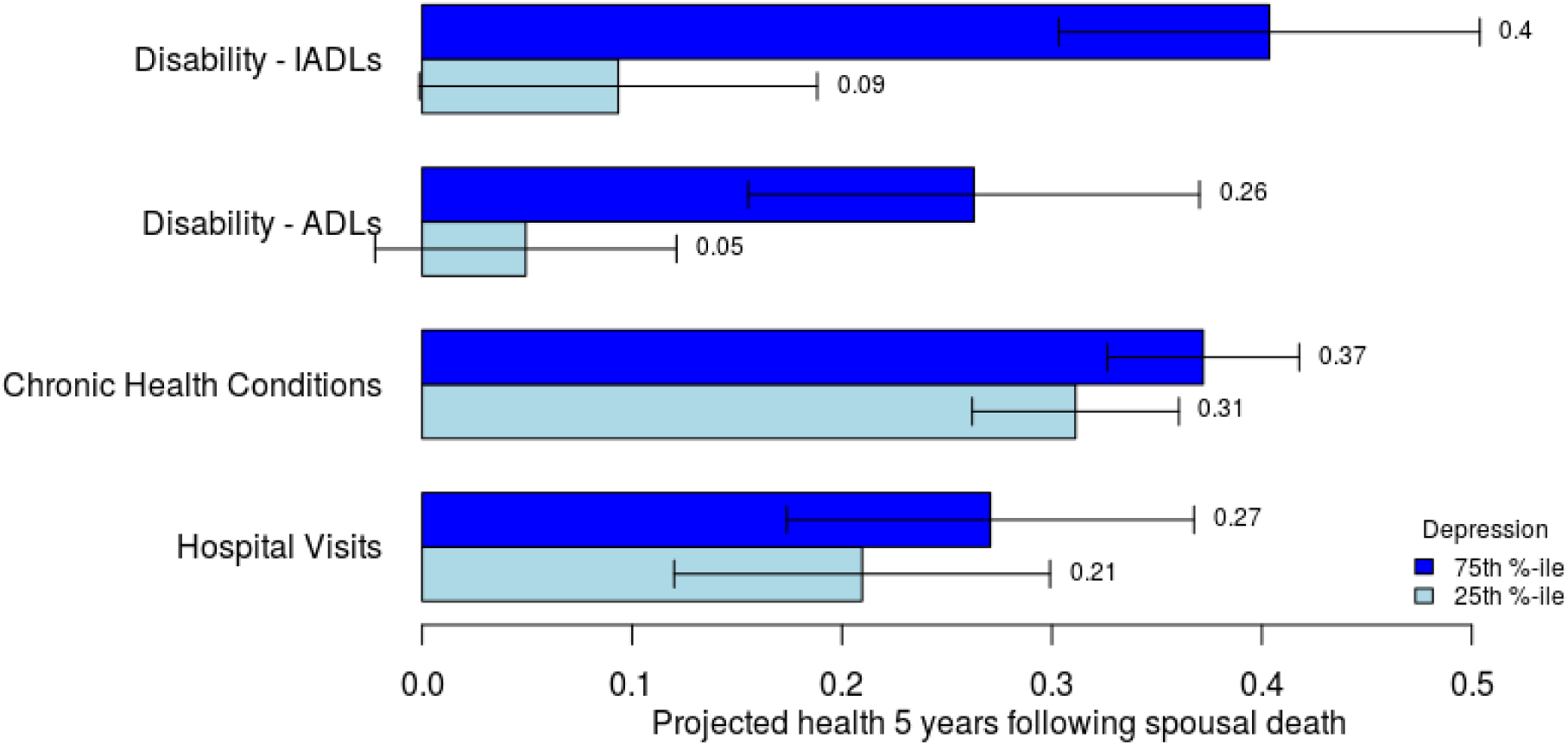
Higher levels of bereavement-related depressive symptoms are associated with poor physical health 5 years following spousal death. Figure graphs predicted values and 95% confidence intervals for physical health outcomes of older participants (aged 74 and older at the time of spousal death) with relatively high levels of bereavement-related depressive symptoms (75^th^ percentile, dark blue bars) and relatively low levels of bereavement-related depressive symptoms (25^th^ percentile, light blue bars). Analysis included all observations within five years of spousal death (minimum N=15159 observations of N=2268 individuals). Health outcomes were adjusted for aging-related decline and period effects (outcome measures were residualized for age (five splines), sex, interactions between the age-based splines and sex, and HRS observation wave). Predicted health values are denominated in HRS standard deviation units. Confidence intervals were estimated via 1000 bootstrap replications. Model results are reported in Supplemental Table S6.

Parallel to analysis of morbidity and disability, older HRS participants who experienced more severe depressive symptoms following their spouse’s deaths were more likely to die themselves over the subsequent 5 years (**Figure 4**). For respondents over 74 at time of spousal death, those with relatively higher levels of bereavement-related depression had a 60% greater risk of death over the next five years as compared to participants with relatively lower levels of bereavement-related depression (for the 75th percentile, risk of death was 16% (95% CI 13-19%); for the 25th percentile, risk of death was 10% (95% CI 7-13%); based on **Supplemental Table S7**).

**Figure 4.**
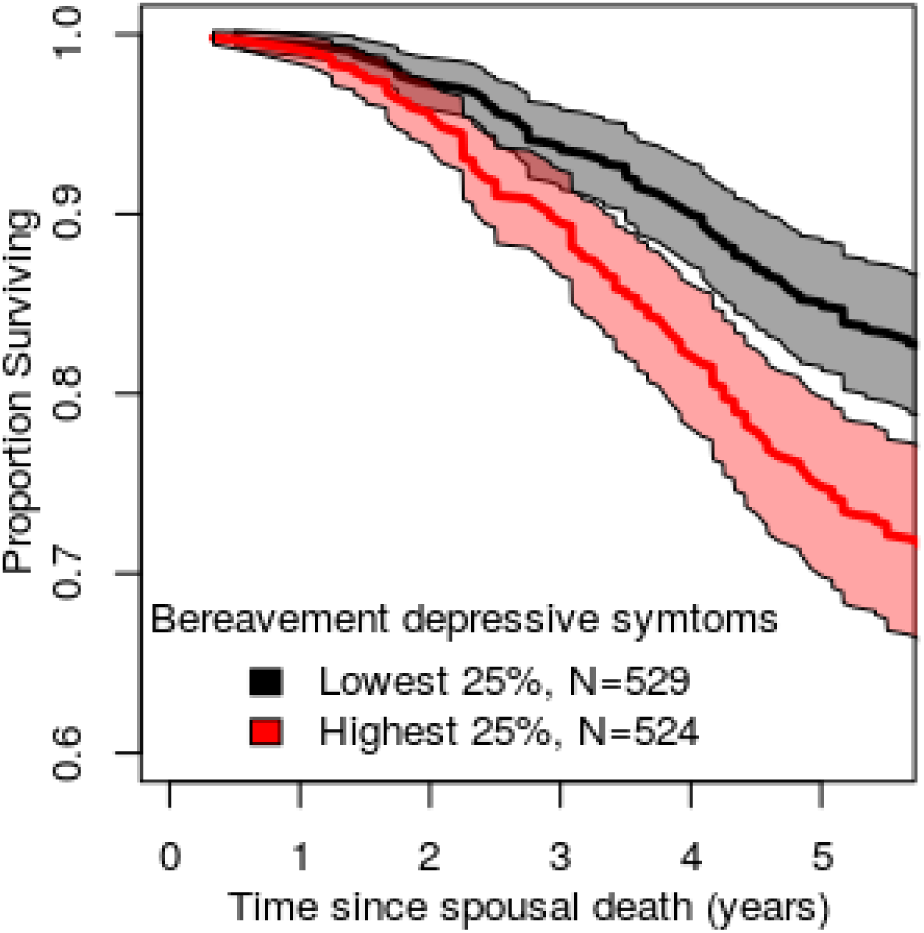
Survival among bereaved participants stratified by level of depression during bereavement. The figure plots survival curves for participants with relatively low levels of bereavement-related depressive symptoms (25^th^ percentile and below, black slope) and relatively high levels of bereavement-related depressive symptoms (75^th^ percentile and above, red slope). The figure shows poorer survival outcomes for those experiencing more severe depressive symptoms in bereavement.

## DISCUSSION

We analyzed mental- and physical-health sequelae of bereavement and connections between these two health outcomes in longitudinal data tracking 4,629 widowed older adults in the US Health and Retirement Study (HRS). We found that widows and widowers who experienced more severe increases in depressive symptoms in the weeks and months following their spouse’s death went on to suffer steeper decline in physical health in the years afterward, even after accounting for health changes during the period leading up to spousal death. This pattern of decline in physical health was more pronounced among participants who were in their 70s and 80s at the time of their spouse’s death as compared to participants who were in their 50s and 60s.

The relationship between depressive symptoms and aging-related physical-health decline is likely to be bidirectional (29,30). Our analysis disentangles the pattern of reciprocal causation to isolate associations of depressive symptoms with subsequent physical health decline through three features of the research design. First, to guard against reverse causation, we tested short-term depressive symptoms as predictors of medium-to-long-term changes in morbidity, disability, and mortality. Second, to rule out confounding by stable individual differences, we analyzed within-person change over time. Third, to rule out confounding by pre-bereavement trends, we used discontinuity analysis to isolate health changes occurring after spousal death. Together, these design features identify a prospective association between depressive symptoms in the immediate aftermath of spousal death and increases in morbidity, disability, and mortality risk over the five years following the death. In sum, our analysis provides new evidence linking mental-health problems with subsequent physical-health decline in later life. These findings have implications for research into connections between mental and physical health in aging and for clinical and public health approaches to bereavement in older adults.

For future research, studies are needed to investigate mechanisms of the prospective association between bereavement-related depression and subsequent physical-health decline. One possibility is that transient symptoms of depression following spousal death induce lasting shifts in behavior that contribute to health decline, such as increased sedentary lifestyle. Such a process could also occur at the molecular level, with depression related changes in immune-system dysregulation and chronic inflammation (31,32) persisting after depressive symptoms resolve. Studies are also needed to test if specific risk factors predispose to bereavement-related health declines. Exposures accumulating from early-life including perinatal insults, childhood adversity, and socioeconomic disadvantage across the life course are associated with increased vulnerability to depression and greater burden of physical-health problems (33– 36). And, as we have previously shown, genetic differences between individuals may explain variation in risk for depression in bereavement (27). Future research should test if individuals at high-risk for bereavement-related depression also experience more pronounced physical health declines following adverse events in aging. Importantly, because these risks can be measured retrospectively, studies do not require prospective assessments of depression in the immediate aftermath of adverse events.

For clinical and public-health approaches to managing bereavement in older adults, our analysis does not establish that treatment of mental health sequelae of bereavement could prevent subsequent physical health decline. However, the association identified in our study can inform prognosis for older adults who have suffered the loss of a spouse. Short-term, transient depressive symptoms related to bereavement are a harbinger of long-term physical health decline in older adults. At minimum, the results reported here amplify calls for increased efforts to integrate mental health screening into geriatrics practice (37,38). They also suggest that follow-up to positive screens should extend beyond efforts to address symptoms of depression and include efforts to monitor and prevent physical health decline. To date evidence for efficacy of depression screening in primary care is limited (39). Findings from this study suggest that evaluating effectiveness of depression screening in geriatric patients should include monitoring medium-term physical health outcomes as well as depressive symptoms.

We acknowledge limitations. The CES-D is an incomplete measurement of mental-health sequelae of bereavement. Replication of findings in studies with comprehensive assessment of depressive symptomatology and evaluation of other symptom dimensions is needed. An advantage of the brief symptom inventory used is that it approximates a tool that could be deployed in clinical practice. All mental- and physical-health sequelae were reported by the participants. Common reporter bias could inflate associations. However, physical-health sequelae continued to worsen years after participants’ mental health had returned to pre-bereavement levels. Thus, associations were not driven by elevated depressive symptoms at the time participants reported physical health problems. Nevertheless, replication of findings in data linking participants with health care claims or clinical exam data will clarify effect-sizes. Observations of HRS participants immediately following their spouse’s death may selectively exclude participants experiencing the most severe mental-health sequelae of bereavement. However, we did not observe a difference in the time intervals for observations bracketing spousal death as compared to other pairs of adjacent observations (**Supplemental Figure S2B**). A related limitation is that mortality selection may conceal the full extent of links between acute depressive symptoms of bereavement and physical health decline. We do not observe participants who die during the interval between their spouse’s death and the next HRS interview. These participants might choose to postpone their HRS interviews. However, we expect such missing data would bias towards estimates toward the null, making our estimates conservative.

With an aging population, the number of older adults at risk for death of their spouse is growing. Our findings suggest that monitoring of depressive symptoms in bereaved older adults may help identify persons at risk for future health decline. In addition, findings suggest new directions for research to investigate mechanisms linking mental and physical health in aging.

## Data Availability

Data are publicly available.

http://hrsonline.isr.umich.edu/

## Acknowledgements

AH is supported by the National Institutes on Aging R01-AG026291. DWB is supported by an Early Career Fellowship from Jacobs Foundation. This work was partially supported by National Institute on Aging grant R21AG054846 and Russell Sage Foundation grant 1810-08987. The Health and Retirement Study is sponsored by the National Institute on Aging (grant number NIA U01AG009740) and is conducted by the University of Michigan.

